# “Association of Poor Housing Conditions with COVID-19 Incidence and Mortality Across US Counties.”

**DOI:** 10.1101/2020.05.28.20116087

**Authors:** Khansa Ahmad, Sebhat Erqou, Nishant Shah, Umair Nazir, Alan Morrison, Gaurav Choudhary, Wen-Chih Wu

## Abstract

**Objective:** Poor housing conditions have been linked with worse health outcomes and infectious spread in communities but its relationship with incidence and mortality of COVID-19 is unknown. Therefore, we undertook this study to determine the association between poor housing condition and COVID-19 incidence and mortality in US counties.

**Methods:** We conducted cross-sectional analysis of county-level data from the US Centers for Disease Control, US Census Bureau and John Hopkins Coronavirus Resource Center for 3141 US counties. The exposure of interest was percentage of households with poor housing conditions (one or more of: overcrowding, high housing cost, incomplete kitchen facilities, or incomplete plumbing facilities). Outcomes were incidence rate ratios (IRR) and mortality rate ratios (MRR) of COVID-19 across US counties through 4/21/2020. Multilevel generalized linear modeling was utilized with adjustment for population density and county characteristics including demographics, income, education, prevalence of medical comorbidities, access to healthcare insurance and emergency rooms, and state-level COVID-19 test density.

**Results:** Across 3135 US counties, the mean percentage of households with poor housing conditions was 14.2% (range 2.7% to 60.2%). The mean (SD) incidence and mortality of COVID-19 were 255.68 (2877.03) cases and 13.90 (272.22) deaths per county, respectively. In the fully adjusted models, with each 5% increase in percent households with poor housing conditions, there was a 50% higher risk of COVID-19 incidence (IRR 1.50, 95% CI: 1.38 – 1.62) and a 42% higher risk of COVID-19 mortality (MRR 1.42, 95% CI: 1.25 – 1.61). Results remained similar using earlier timepoints (3/31/2020 and 4/10/2020).

**Conclusions and Relevance:** Counties with a higher percentage of households with poor housing had higher incidence of, and mortality associated with, COVID-19. These findings suggest targeted health policies to support individuals living in poor housing conditions should be considered in further efforts to mitigate adverse outcomes associated with COVID-19.

## Introduction

Coronavirus disease-2019 (COVID-19) is a rapidly evolving pandemic caused by the novel enveloped RNA beta-coronavirus, Severe Acute Respiratory Syndrome Coronavirus 2 (SARS-CoV-2)(1). As of April 26, 2020, SARS-COV-2 has infected over 2 million people worldwide and led to >200k deaths,^(2)^. The infectivity potential of SARS-CoV-2 is a function of its high basic reproductive number (R_0_) which has been estimated to be from 2.2 to as high as 5.7, as compared to R_0_ of 1.3 for influenza virus(3–5). Given the high infectivity and lack of a vaccine, one of the important mitigation strategies employed worldwide in order to decrease the rate of spread and ‘flatten’ the infection curve has been social and physical distancing(3, 6). However, physical distancing can be compromised if the living environment for various reasons, such as poor housing conditions, prevents the individuals to stay isolated. Problems associated with poor housing include overcrowding, high housing cost, lack of kitchen facilities or lack of plumbing facilities. Given the overcrowding and the inherent need to utilize communal facilities (bathrooms or kitchens), physical distancing would likely be compromised in the households experiencing poor housing conditions. Poor housing conditions have already been linked to an increased spread of respiratory infections(7). Furthermore, there is robust literature linking poor housing conditions to worse health outcomes(8–11). Given that a common mechanism of SARS-CoV-2 transmission is through respiratory aerosols and droplet contact, poor housing conditions can potentially facilitate the spread of the infection(12). Since the relationship between poor housing conditions and COVID-19 outcomes are currently unknown, we undertook a study to test the hypothesis that the percentage of households with poor housing conditions in US counties are associated with higher incidence and mortality of COVID-19 across 3135 counties in the United States (US).

## Materials and Methods

We conducted a cross sectional ecological analysis of the 3141 US counties using publicly available data relating poor housing conditions with COVID-19 outcomes. Counties with missing data for poor housing condition (n = 6) were excluded yielding a sample size of 3135 US counties. These counties were Prince of Wales-Outer Ketchikan, Skagway-Hoonah-Angoon, Wrangell-Petersburg and Kusilvak from Alaska, Bedford City from Virginia and Oglala Lakota from South Dakota. Counties in US territories (American Samoa, Guam, Northern Mariana Island, Puerto Rico and US Virgin Islands) were not included in our analysis. Since there is no individual identifying information, the data were in aggregate by county, and publicly available from the Centers for Disease Control (CDC), US census Bureau and John Hopkins Coronavirus Resource Center,(2) the protocol received exemption from the Providence Veterans Affairs Medical Center Institutional Review Board(2, 13, 14).

### Exposure

The exposure of interest was the percentage of households in a county with poor housing conditions which has been published as percentage of households with severe housing problems (2010 – 2014) by the CDC(13). These households were identified as having any of the four problems: 1) overcrowding, 2) high housing cost burden, 3) incomplete kitchen facilities, and 4) incomplete plumbing facilities. In further breakdown, overcrowding is defined as more than one person per room, high housing cost as more than 50% of the monthly household income allocated towards housing cost (including utilities), incomplete kitchen facilities as lacking a sink with running water, stove or range, or a refrigerator, and incomplete plumbing facilities as lacking of hot and cold piped water, a flush toilet, or a bathtub/shower(15). To facilitate interpretation, we studied the poor housing conditions as a continuous variable by units of 5% increase in households with poor housing conditions, which is roughly equivalent to the standard deviation (SD) of the variable. Additionally, we also categorized the counties according to approximate quartiles based on percentage of households with poor housing conditions (rounded values were used as cut off).

### Outcome

The outcomes of interest were incident COVID-19 cases and COVID-19 deaths per county. We obtained the COVID-19 incidence and mortality data from the John Hopkins Coronavirus Resource Center, where it has been published for “public health, educational, and academic research purposes.”(2) We obtained the cumulative data for three dates of 10-day intervals: March 31^st^, April 10^th^ and April 21^st^, which allowed us to test the robustness of our findings across three temporal cross sections of the reported data. April 21^st^ data were utilized as the main analysis.

### Covariates

We obtained data on county level variables related to COVID-19 spread and outcomes based on literature. The data regarding total population (2010) and population density (population per square miles of land area of a county) were obtained to account for the exposure pool(13). In order to account for socioeconomic disparity, data on median household income of a county (2016) and percentage of residents without a high school diploma (2013–2017) were included(13).

County demographic data was collected because male sex, older age and percent of racial minority have been linked to a higher risk of, and mortality in, COVID-19(13, 16–18). This included percentage of male residents (2010), median age (2010), and percentage of white, black, Hispanic or Latino, Asian, Native Hawaiian or Pacific Islander; and American Indian or Alaska Native residents (2013 – 2017)(13).

From the very initial reports, COVID-19 was seen to drastically effect individuals with a heavier burden of comorbidities(16, 17). We therefore collected data for percentage of residents diagnosed with diabetes and those with obesity (2015)(13). Furthermore, Medicare hospitalization data for: hypertension, ischemic stroke, myocardial ischemia, heart failure and dysrhythmia (2014 – 2016) were obtained as a surrogate for cardiovascular disease burden in the county(13).

The morbidity and mortality associated with COVID-19 has been linked to respiratory failure(19). We therefore collected variables that affect respiratory health: annual concentration of particulate matter 2.5 μ (pm2.5) (2014) and percentage of residents who are current smokers (2017)(13).

We accounted for the percentage of adults without health insurance under 65 years (2016) and number of hospitals with emergency rooms (ER) (2016) in each county, as surrogates of access to care. The number of hospitals may also influence the number of cases detected in the county. Furthermore, we obtained the total number of tests conducted per state (cumulative up to April 21^st^) and calculated the ratio of tests to the total population of the state (test density)(20).

### Statistical Analysis

County covariates were described as mean ± SD and range for continuous variables and as number (%) for categorical variables. Linear regression was used to assess linear trend of the county covariates across the four quartiles.

We graphed the correlation between poor housing and number of COVID-19 cases and COVID-19 deaths. Given non-normal distribution, we used log transformation (log × +1) of the exposure and outcome variables for the scatter plot and calculated the Pearson’s correlation coefficient.

We utilized generalized linear multilevel models with a negative binomial distribution family and a log link function to study the association between the relative risk of each 5% change in households with poor housing conditions and the incidence (first analysis) and mortality (second analysis) of COVID-19, with total population of a county as denominator. To account for clustering effect due to poliy, social and behavioral similarities across counties within the same state, we applied a random intercept for the state. The covariance matrix was specified as unstructured. We adjusted for different categories of variables in the regression model in a stepwise fashion: 1) population density and test density, 2) demographics (% male, median age,% white), 3) socioeconomic status (median household income, % residents with lack of high school education), 4) respiratory exposure (annual ambient PM2.5, % current smokers), 5) prevalence of comorbidities (% diagnosed with diabetes, % diagnosed with obesity), 6) Medicare hospitalization rates (hypertension, ischemic stroke, myocardial ischemia, heart failure, dysrhythmia), and 7) Access to healthcare (% adults without health insurance under 65 years and number of hospitals with ER). The fully adjusted model included all the aforementioned variables. We reported incidence rate ratios (IRR) and mortality rate ratios (MRR), respectively, which are interpretable as the relative increase in incidence and mortality for COVID19, respectively, for each 5% increase in households with poor housing conditions.

We conducted several sensitivity analyses to assess the robustness of our findings. 1) In lieu of percent white population in a county, we replaced it with the percent breakdown of the minority population: black, Hispanic or Latino, Asian or Native Hawaiian or Pacific Islander and American Indian or Alaska Native residents per county in the fully adjusted model. 2) Quartile Analysis – we also studied quartiles of percent households with poor housing conditions. We tested the quartiles in the fully adjusted model as an ordinal variable (i.e. linear effect across quartiles) and as categorical (dummy) variables (using 1^st^ quartile as referent). 3) Temporal Analysis – Since the main analysis is using the most recent data (April 21^st^, 2020), we repeated our analyses in two earlier time points for COVID-19 incidence and mortality as outcomes, on March 31^st^ and April 10^th^, to account for and understand temporal changes, if any, in this association.

A two-sided p value of < 0.05 was considered statistically significant. All analyses were conducted in Stata SE statistical software (Stata Corp, Texas, v. 15.0).

## Results

Across 3135 US counties, the mean (range) percentage of households with poor housing conditions was 14.2% (2.7% to 60.2%). The modified quartiles of percentage of households with poor housing conditions were 2.7% to 11% (Quartile1), 11.1% to 14.0% (Quartile2), 14.1% to 17.0% (Quartile3) and

17.1% to 60.2% (Quartile4). Till April 21^st^, there were a total of 144190 confirmed COVID-19 cases assigned to these 3135 US counties and 14887 COVID-19 deaths. The mean (SD) incidence and mortality of COVID-19 were 255.68 (2877.03) cases and 13.90 (272.22) deaths per county, respectively. Table 1 describes characteristics of counties across the US, overall and stratified by quartiles of percentage of households with poor housing conditions.

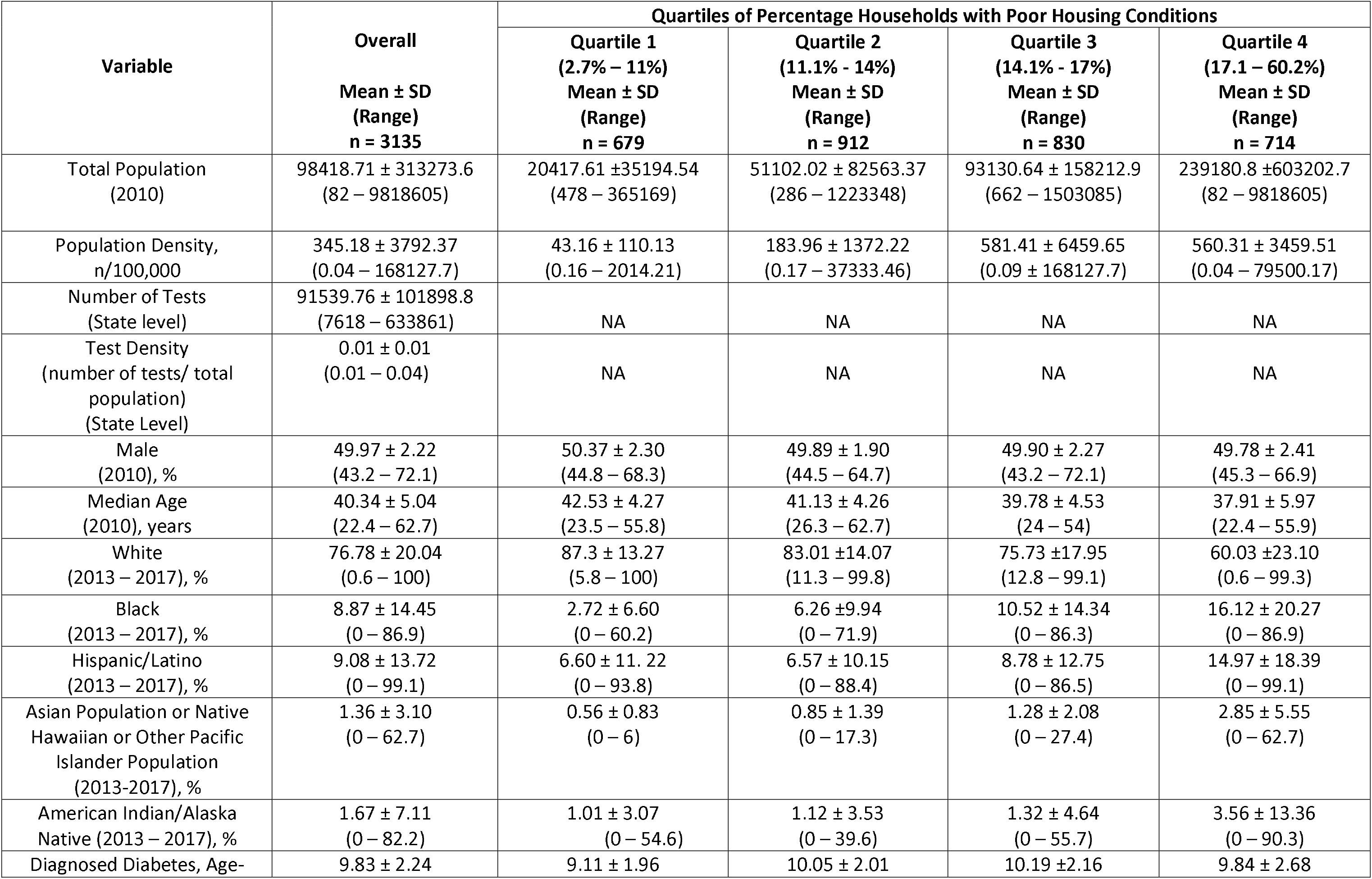
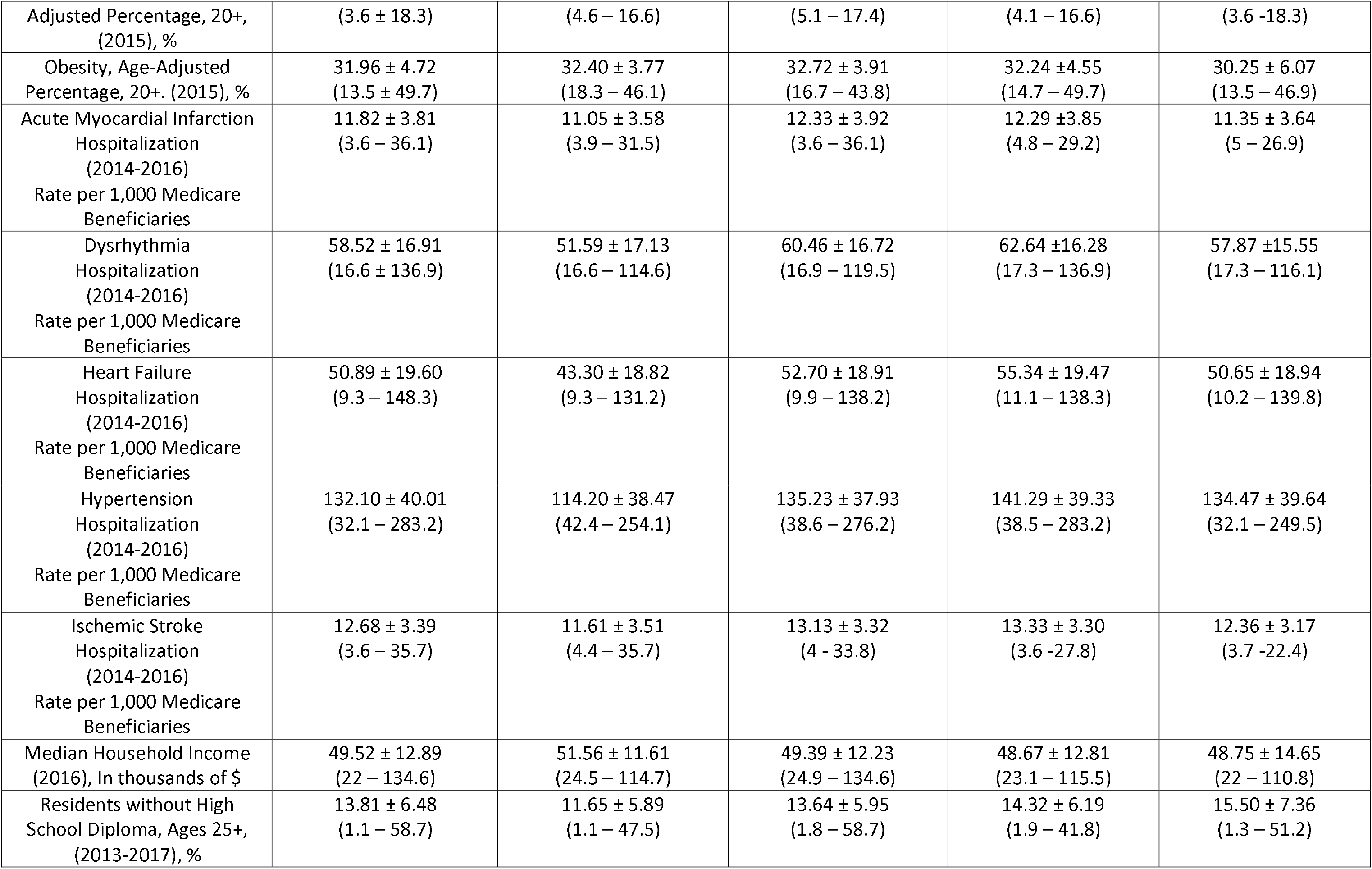
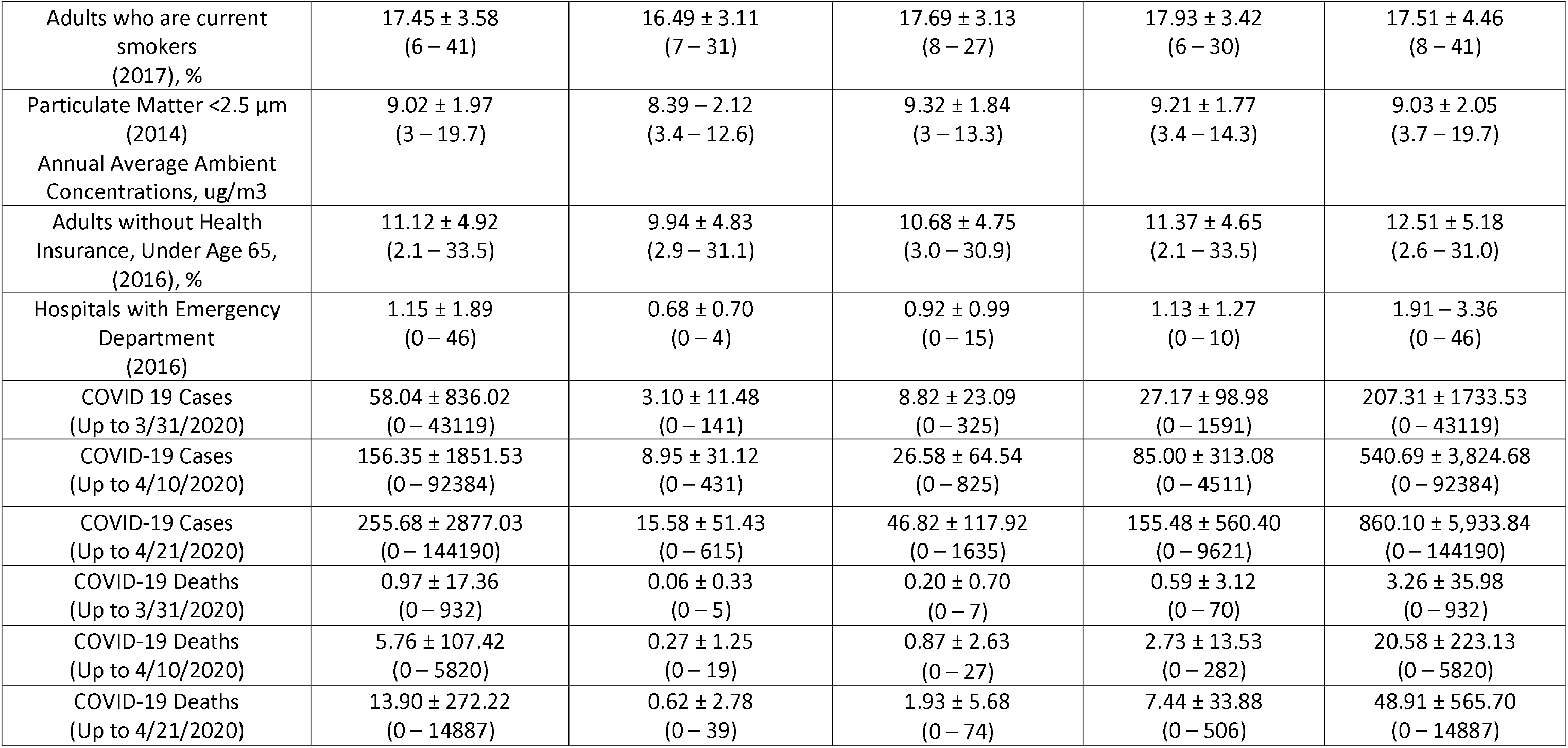

The mean number of COVID-19 cases and deaths for all time points, March 31^st^, April 10^th^ and April 21^st^ increased across increasing quartiles of percentage of households with poor housing conditions (all p’s <0.001). Similarly, increasing quartiles of percent households with poor housing conditions were associated with higher county population, population density, percentage of minority residents, percentage of residents without high school diploma, prevalence of diabetes, Medicare hospitalization rates for hypertension, ischemic stroke, heart failure and dysrhythmia, percentage of current smokers, annual PM2.5 levels, percentage of adults < 65 years without health insurance and number of hospitals with ER facilities (all p’s <0.001). Conversely, a decrease in the median household income and prevalence of obesity was observed across increasing quartiles of percent households with poor housing conditions (both p’s <0.001).

There was a moderately strong correlation between the log-transformed percentage of households with poor housing conditions and log-transformed number of COVID-19 cases (r = 0.44, p < 0.001) and COVID-19 deaths (r = 0.47, p<0.001) across US counties (**Figure 1**).

**Figure 1a-b:**
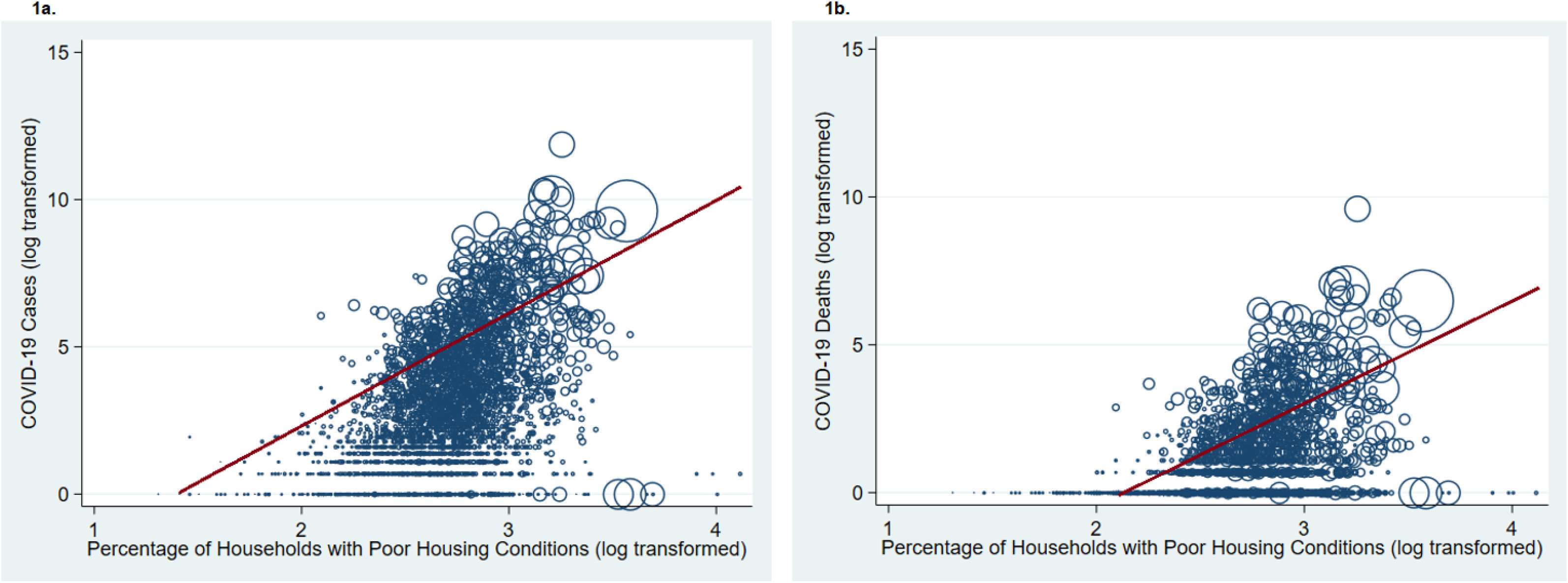
Scatter plot of COVID-19 Cases & Deaths by percentage of Households with Poor Housing Conditions across 3135 US counties.

Using the COVID-19 data from April 21^st^, we found that for each 5% increase in poor housing condition per county, there is a 59% increase in the relative risk of COVID-19 incidence (IRR 1.59, 95% confidence interval [CI]: 1.49–1.70) (**Table 2**). The association was only slightly attenuated after adjustment for an extensive list of county covariates (**Table 2**). The IRR for the fully adjusted model (Model VIII) was 1.50 (95% CI: 1.38 – 1.62) (**Table 2**). Secondary analyses categorizing the exposure into quartiles (**Figure 2a**) or using data from March 31^st^ and April 10^th^ yielded comparable results (**Table 2**).

**Table 2:**
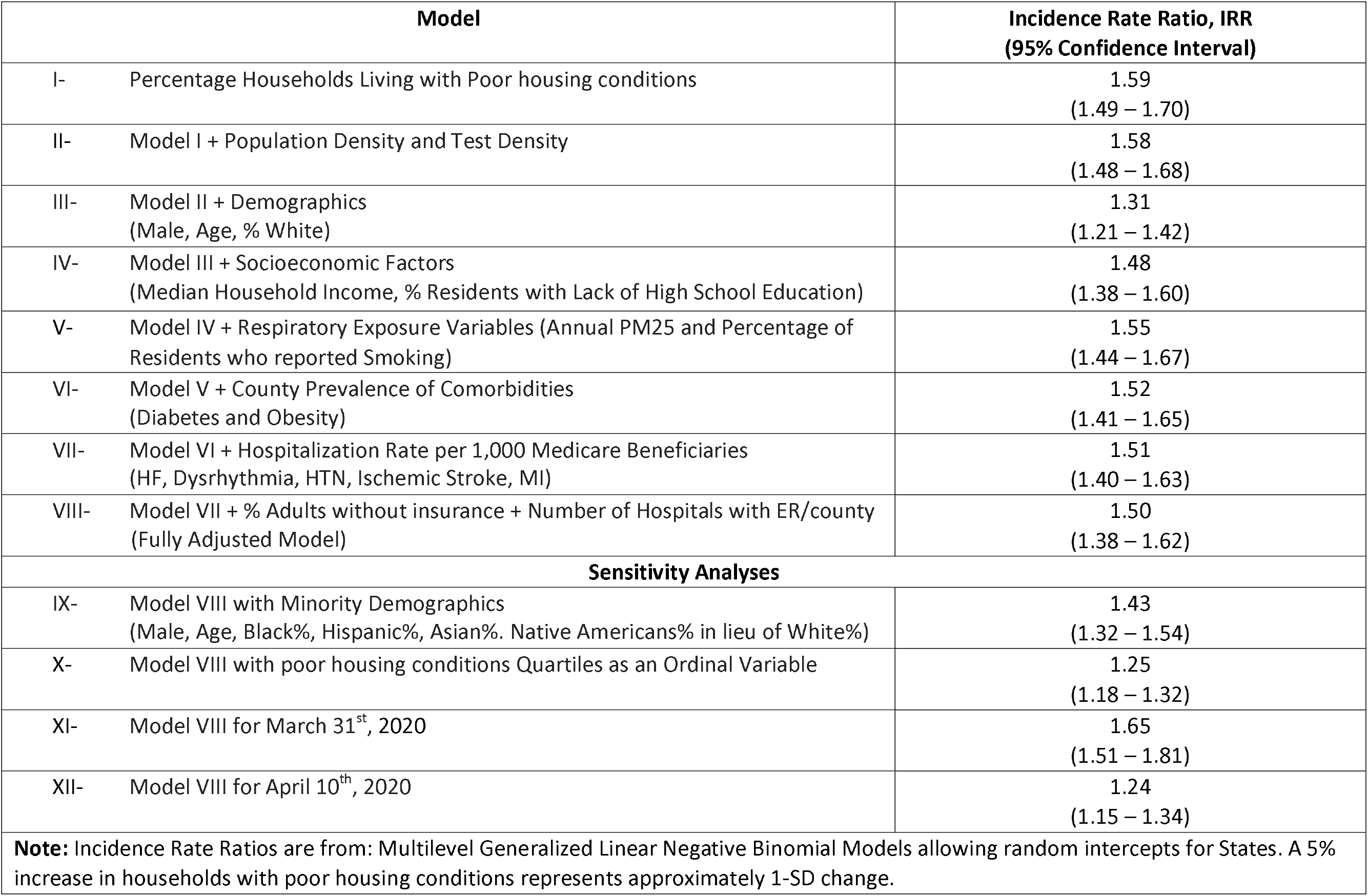
Association of county Covid-19 incidence as of April 21, 2020 with percent households with poor housing conditions.

| Model |  | Incidence Rate Ratio, IRR<br>(95% Confidence Interval) |
| --- | --- | --- |
| I- | Percentage Households Living with Poor housing conditions | 1.59<br>(1.49 – 1.70) |
| II- | Model I + Population Density and Test Density | 1.58<br>(1.48 – 1.68) |
| III- | Model II + Demographics<br>(Male, Age, % White) | 1.31<br>(1.21 – 1.42) |
| IV- | Model III + Socioeconomic Factors<br>(Median Household Income, % Residents with Lack of High School Education) | 1.48<br>(1.38 – 1.60) |
| V- | Model IV + Respiratory Exposure Variables (Annual PM25 and Percentage of Residents who reported Smoking) | 1.55<br>(1.44 – 1.67) |
| VI- | Model V + County Prevalence of Comorbidities<br>(Diabetes and Obesity) | 1.52<br>(1.41 – 1.65) |
| VII- | Model VI + Hospitalization Rate per 1,000 Medicare Beneficiaries<br>(HF, Dysrhythmia, HTN, Ischemic Stroke, MI) | 1.51<br>(1.40 – 1.63) |
| VIII- | Model VII + % Adults without insurance + Number of Hospitals with ER/county<br>(Fully Adjusted Model) | 1.50<br>(1.38 – 1.62) |
| **Sensitivity Analyses** |  |  |
| IX- | Model VIII with Minority Demographics<br>(Male, Age, Black%, Hispanic%, Asian%, Native Americans% in lieu of White%) | 1.43<br>(1.32 – 1.54) |
| X- | Model VIII with poor housing conditions Quartiles as an Ordinal Variable | 1.25<br>(1.18 – 1.32) |
| XI- | Model VIII for March 31<sup>st</sup>, 2020 | 1.65<br>(1.51 – 1.81) |
| XII- | Model VIII for April 10<sup>th</sup>, 2020 | 1.24<br>(1.15 – 1.34) |
| **Note:** Incidence Rate Ratios are from: Multilevel Generalized Linear Negative Binomial Models allowing random intercepts for States. A 5% increase in households with poor housing conditions represents approximately 1-SD change. |  |  |

**Figure 2a-b:**
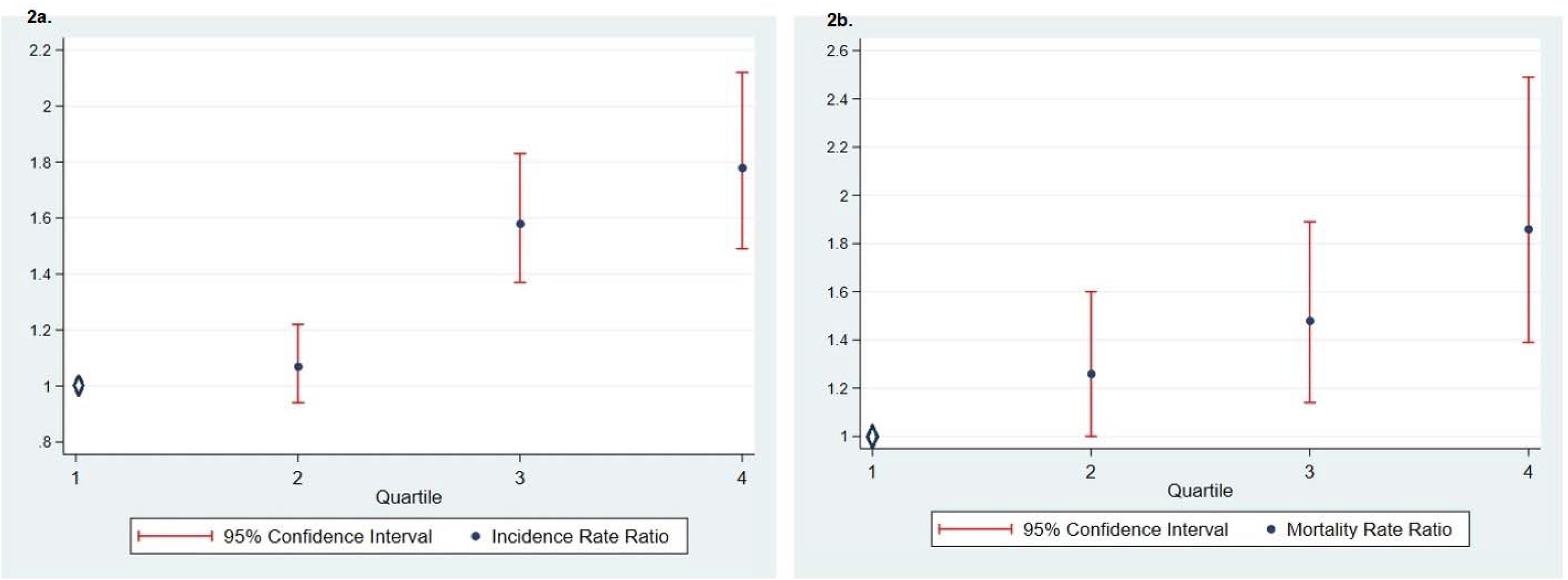
Relative Risk Increase in Incidence and Mortality of Covid-19 with Each Quartile of Percentage of Households with Poor housing conditions.

Similarly, using the COVID-19 data from April 21^st^, we found a 63% increase in the relative risk of COVID-19 mortality for each 5% increase in poor housing condition per county, (MRR 1.63, 95% CI: 1.48 – 1.79) (Table 3). The association was only mildly attenuated after adjustment (Table 3), and remained highly significant in the fully adjusted model (Model VIII), with an MRR of 1.42 (95% CI 1.25 – 1.61) (Table 3). Secondary analyses categorizing the exposure into quartiles (Figure 2b) or using data from March 31^st^ and April 10^th^ yielded comparable results (Table 3).

**Table 3:** Association of county Covid-19 Deaths as of April 21, 2020 with percent households with poor housing conditions.

| Model |  | Mortality Rate Ratio, MRR<br>(95% Confidence Interval) |
| --- | --- | --- |
| I- | Percentage Households Living with Poor housing conditions | 1.63<br>(1.48 – 1.79) |
| II- | Model I + Population Density and Test Density | 1.62<br>(1.47 – 1.78) |
| III- | Model II + Demographics<br>(Male, Age, White%) | 1.25<br>(1.12 – 1.40) |
| IV- | Model III + Socioeconomic Factors<br>(Median Household Income, % Residents with Lack of High School Education) | 1.41<br>(1.26 – 1.58) |
| V- | Model IV + Respiratory Exposure Variables (Annual PM25 and Percentage of Residents who reported Smoking) | 1.49<br>(1.32 – 1.67) |
| VI- | Model V + County Prevalence of Comorbidities<br>(Diabetes and Obesity) | 1.44<br>(1.27 – 1.63) |
| VII- | Model VI + Hospitalization Rate per 1,000 Medicare Beneficiaries<br>(HF, Dysrhythmia, HTN, Ischemic Stroke, MI) | 1.44<br>(1.27 – 1.64) |
| VIII- | Model VII + %Adults without insurance + Number of Hospitals with ER/county<br>(Fully Adjusted Model) | 1.42<br>(1.25 – 1.61) |
| **Sensitivity Analyses** |  |  |
| IX- | Model VIII with Minority Demographics<br>(Male, Age, Black%, Hispanic%, Asian%, Native Americans% in lieu of White%) | 1.37<br>(1.21 – 1.56) |
| X- | Model VIII with poor housing conditions Quartiles as an Ordinal Variable | 1.22<br>(1.11 – 1.34) |
| XI- | Model VIII for March 31<sup>st</sup> | 1.36<br>(1.10 – 1.67) |
| XII- | Model VIII for April 10th | 1.31<br>(1.14 – 1.51) |
| Note: Mortality Rate Ratios are from: Multilevel Generalized Linear Negative Binomial Models allowing random intercepts for States. A 5% increase in percent households with poor housing conditions represents approximately 1-SD change. |  |  |

## Discussion

To our knowledge, this is the first nationwide study to investigate county level association of COVID-19 incidence and mortality with percentage of households facing poor housing conditions in the US. Our study showed that with each 5% increase in percent households with poor housing conditions, there was a 50% higher risk of COVID-19 incidence and a 42% higher risk of COVID-19 mortality across US counties. Findings remained similar in three different time points and after accounting for county-level population density and state test density, demographics, socioeconomic status, prevalence of comorbidities, respiratory exposure, lack of health insurance and number of ER facilities.

Of the four issues categorized under poor housing conditions, overcrowding offers the most direct explanation for the higher incidence and mortality of COVID-19. Overcrowding has been associated with spread of respiratory illnesses like tuberculosis and influenza which have aerosol and droplet transmissions, both of which are potential modes of transmission for COVID-19(7, 12, 21–23). The World Health Organization (WHO) published housing and health guidelines in 2018 identifying poor housing related environmental risk factors including overcrowding, air and water quality and lack of access to adequate plumbing and sanitation, as factors contributing to the burden of infectious diseases including airborne respiratory illnesses(24). The 2003 spread of severe acute respiratory syndrome (SARS) epidemic in Hong Kong was worst in the Amoy Garden estate which was overcrowded and had significant plumbing and sanitation problems(25). An initial study from China investigating the initial COVID-19 outbreaks also showed that 79.9% of outbreaks occurred indoors, almost all in apartment settings(26). Evidence from the Influenza epidemic of 1918 showed not only increased spread but also increased severity of the disease as a result of overcrowding(27). Overcrowding associated with a more severe infectious disease process may offer a potential explanation of the higher mortality associated with poor housing, as overcrowding leads to repeated exposure and potentially a higher viral inoculum. High viral load has been linked to worse COVID-19 clinical outcomes(28–30).

Another feature of poor housing conditions is high cost burden, with more than 50% of the household income going towards housing cost(15). Prior studies have shown an association between high housing costs and delays in seeking healthcare(8, 31). Our study showed that counties with the highest percentage of residents with poor housing conditions also had the lowest median household income and highest percentage of residents without a high school diploma. There are several news reports indicating that minimum wage workers have found it difficult to follow the stay-at-home guidelines, due to a lack of economic resources and being part of essential work force that is mostly an exception to stay-at-home guidelines(32–34). However, an important feature of our study is that although linked to economic status, the associations between poor housing conditions and COVID-19 incidence, and mortality, were independent of the racial composition, median household income, lack of education and lack of health insurance, as the relative risk was only mildly changed and remained highly significant after adjusting for these factors.

A lack of appropriate plumbing and kitchen facilities is also part of poor housing conditions. Availability of appropriate plumbing facilities and in-home clean water has been linked to a decline in infectious disease including lower respiratory tract infections(24, 35). Lack of appropriate facilities interferes with the ability to practice good hand hygiene. Furthermore, lack of these facilities would require the residents to use communal facilities, thereby increasing social contact. It is important to note that SARS-CoV-2 is detectable for up to 72 hours on plastic and stainless steel materials, whereas influenza is detectable for 24–48 hrs(36, 37). This poses another public health risk for transmission to other individuals sharing a crowded space, in the absence of meticulous hygiene.

Our findings have health policy implications as they identify a particularly vulnerable population to be at heightened risk and also the potential pathways for public health interventions during the current COVID-19 pandemic. In addition, our study adds to a robust body of evidence for other disease processes, which has shown that inadequate housing is a public health hazard especially in relation to infectious diseases and highlights the importance of finding short (e.g. better access to clean water and bathrooms) and long-term (e.g. overcrowding, cost) solutions to problems surrounding poor housing to help contain or mitigate the spread of COVID-19. Health education via print or social media targeted at the population at risk to improve awareness of preventative measures and hygiene, establishment of hygiene protocols and increased availability of mobile bathrooms and cleaning supplies at communal facilities can be considered to help mitigate the COVID-19 spread(38, 39).

The strength of this study is that this is a nationwide report of 3135 counties across US, which allows for a large sample size and generalizability of our findings. Furthermore, to our knowledge, this is the first study to establish an association between the incidence and mortality of COVID-19 and poor housing conditions. The limitations of the study also merit consideration. First, the county-level covariate data utilized were from earlier time period, and hence may have weakened the strength of the associations. However, we utilized the most updated results publicly available. Furthermore, the assumption that county age structure and ethnic composition does not quickly change over the span of several years is the current approach shared by the US Census methodology (every 10 years). However, the consistent results after accounting for extensive list of covariates and various sensitivity analyses, support the robustness of the findings. Due to limitations of the data, we could not separate the distinct elements (e.g. overcrowding, cost, plumbing, kitchen) that comprised poor housing for better understanding of the problem and targeting of policies. This is also a cross-sectional ecological analysis and does not lend itself to causal inference. Finally, despite careful adjustments and inclusion of covariates, residual confounding cannot be excluded.

## Conclusion

In a nationwide analysis of US county-level data, counties with a higher percentage of households with poor housing had higher incidence of, and mortality associated with, COVID-19. These findings suggest targeted health policies to support individuals living in poor housing conditions should be considered in further efforts to mitigate adverse outcomes associated with COVID-19.

## Data Availability

All data files are available from the John Hopkins COnsortium CDC and census bureau database.

## Disclosures

None

## Acknowledgement

The views expressed in this paper represent the authors and not the Department of the Veterans Affairs.

## Notes

### Competing Interest Statement

The authors have declared no competing interest.

### Funding Statement

no funds

### Author Declarations

The protocol received exemption from the Providence Veterans Affairs Medical Center Institutional Review Board.

